# A health equity perspective on data-driven treatment decisions in cardiovascular care: risk assessments versus individualized treatment rules

**DOI:** 10.1101/2025.09.08.25335035

**Authors:** Safiya Sirota, Norrina B. Allen, R. Graham Barr, Daniel Malinsky

**Affiliations:** Department of Biostatistics, Columbia University; Department of Preventive Medicine, Northwestern University; Departments of Medicine and Epidemiology, Columbia University

## Abstract

**Background:** Medical treatment decisions are often based on estimated global risk scores. When heterogeneity in treatment effects exists, assigning treatment according to estimated individualized treatment rules (ITRs) instead has the potential to improve mean outcomes. To investigate racial and ethnic group differences in treatment rates when comparing antihypertensive medication recommendations from an estimated ITR with a risk score approach.

**Methods:** Data were simulated to emulate observational data with underlying treatment effect heterogeneity in survival times. An ITR and risk score approach were compared to illustrate how the resulting recommendations may disagree. An ITR for prescribing antihypertensives was estimated from 3,281 adults from the Multi-Ethnic Study of Atherosclerosis (MESA), an observational longitudinal cohort study, and compared to the risk-based approach recommended by cardiovascular care guidelines. Hypothetical treatment rates under each “rule” were computed. In the simulation study, the proportion of individuals treated optimally under each rule was calculated. Using MESA, a Chi-square test of independence was performed to determine whether treatment rates differed across racial and ethnic groups.

**Results:** Two benefits of ITRs were shown: they (1) maximize expected survival times and (2) may mitigate racial disparities when treatment effect heterogeneity is expected. Using MESA, the ITR recommended treatment to more participants than the risk score approach across all racial and ethnic groups. A Chi-square test suggested that treatment rates for different “rules” differed significantly across racial and ethnic groups (p < .001).

**Conclusion:** Treatment recommendations varied substantially when assigning treatment using an ITR versus a risk-based approach.

## Introduction

Cardiovascular risk scores such as those based upon the Pooled Cohort Equations (PCEs) are used to guide treatment initiation and intensification using an individual’s global atherosclerotic cardiovascular disease (ASCVD) risk.^1–4^ There exist several alternative risk scores derived from various cohorts for predicting CVD outcomes, e.g., the Framingham risk score, Reynold’s risk score, and Multi-Ethnic Study of Atherosclerosis (MESA) risk score.^5^ More recent models, such as the PREVENT models, remove race and ethnicity as a variable in an effort to debias clinical risk prediction and more equitably guide treatment decisions.^6^ All these model-based predictions of clinically relevant outcomes have been recommended to guide patients and providers as they decide on treatment strategies. As algorithmic tools become more sophisticated and AI technologies become more ubiquitous in clinical settings, the practice of informing decisions using risk assessment instruments is likely to grow. However, there is a concern that risk assessment tools used to make decisions may introduce or exacerbate health disparities. For example, when subgroups of the population are underrepresented in the data, risk predictions for these groups may be poor. This is troubling especially in cases where an accurate prediction of high risk is required for the receipt of some medical procedure that can only be allocated to a small number of people.^7^ In settings where resources are unconstrained and data are high-quality and representative, such as for the risk prediction of cardiovascular events, the health equity consequences of using risk scores for treatment decisions have not been rigorously investigated when underlying effect heterogeneity is present. In this case, accurate risk score predictions may not be sufficient to inform appropriate treatment decisions to reduce disease occurrence in all subpopulations, since the treatment may be less effective for some and more effective for others.

There is a large and growing technical literature on individualized treatment rules (ITRs) that combines insights from statistics, computer science, and causal modeling to determine “optimal” treatment strategies for patients based on their individual characteristics.^8,9^ Optimality is typically defined as minimizing expected mortality or chance of adverse medical events. Conditional average treatment effects are estimated and used to allocate treatments optimally, rather than indirectly based on estimated risk categories. ITRs have been used to support treatment decisions in some clinical settings because when accurate estimation is feasible, they can out-perform risk-based treatment assignments, reducing the burden of disease.^10^ These rules also enjoy provable mathematical guarantees when models are correctly specified.^8,9^ The difference between these two approaches manifests when there exists heterogeneity in treatment effects, as visualized in Figure 1. Some subjects may be essentially non-responsive to treatment, even if they are at high baseline risk; the key is to identify subjects that are likely to decrease their relative risk of adverse outcomes under treatment (relative to no treatment), regardless of their position on the distribution of baseline risk. ITRs may provide an approach to directly modeling “optimal” treatment choices but have not been incorporated into CVD prevention and treatment guidelines. This idea also underlies the Predicted Approaches to Treatment effect Heterogeneity (PATH) guidelines, and researchers have begun to explore the benefits of using ITRs estimated from RCT data using machine learning to improve health outcomes.^11,12^

**Figure 1:**
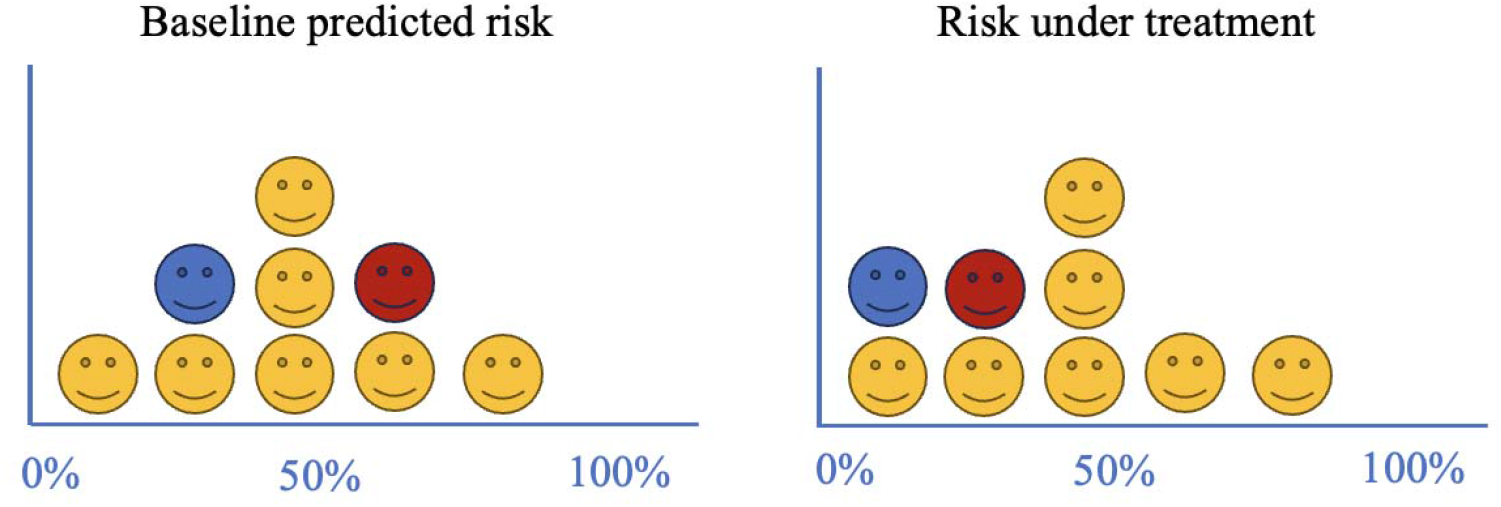
An illustration of hypothetical treatment effect heterogeneity. Left panel: a histogram of baseline predicted risk for a CVD event. Right panel: the risk of a CVD event under the counterfactual that all participants are prescribed treatment. Here, all yellow participants are not affected by treatment, even the one with the highest estimated risk. However, both the blue and red subjects, who may be at moderate or higher risk to begin with, see improvements in their risk under treatment.

Our hypothesis is that ITRs may also be superior from an equity perspective. Reliance on estimated baseline risk may lead to disparities in treatment assignment if racial and ethnic group membership is associated with risk factors, but the true treatment effect is heterogeneous in the population. ITRs would instead assign treatments to those who stand to benefit from them. The racial and ethnic diversity of the MESA sample will allow the ITR to assign treatment partially based on these characteristics, if incorporating this information proves useful for improving expected event-free survival time. The induced medication use disparity will be contrasted between a “rule” that recommends medication for every patient at “high risk” of CVD events, versus another “rule” that assigns medication based on an ITR, accounting for treatment effect heterogeneity.

## Methods

### Simulation Study

First, an analysis of simulated data was conducted to illustrate how recommendations determined by an ITR may differ from recommendations based on a risk assessment approach. Data were synthetic and created to emulate what would be accessible from a longitudinal prospective cohort study aimed at evaluating the efficacy of a preventative treatment. Control over the data-generating process, and thus knowledge of true counterfactual outcomes, was necessary to demonstrate that the ITR (1) correctly identified which patients would benefit from treatment and (2) identified the optimal treatment for individuals at the same rate across subgroups.

Throughout, the “optimal” treatment refers to the treatment that maximizes the mean overall survival time until a CVD event i.e., the event-free survival time, given the outcomes of interest are CVD events and not deaths.

#### Data generation

The simulated dataset (n = 10,000) contained for each hypothetical “subject” information on a continuous biomarker *X*, representing blood pressure or cholesterol level, a binary treatment assignment *A*, representing antihypertensive treatment, a binary demographic group indicator *S*, representing demographic group membership (e.g., “majority” vs. “minority” in a setting with just two groups), an event indicator, a survival time, and a binary covariate *Z*. The covariate *Z* represented the unmeasured heterogeneity in the treatment effect across individuals. Treatment was only efficacious for subjects with *Z*=0; any subject with *Z*=1 did not benefit from treatment. Though *Z* was generated and known by us in the simulation, it was excluded from the models described below and thereby represented a factor that was unknown at the time of determination of treatment efficacy. Therefore, *Z* represented an unknown indicator of treatment efficacy that we may assume exists in the real world. The demographic composition of the data was unbalanced, i.e., 70% of the sample comprised the majority *S* group and 30% comprised the minority *S* group. The data reflected group-level differences, specifically that minority participants were less likely to receive treatment historically (OR=0.69, 95% CI: [0.63, 0.75]), representing historical racial bias through undertreatment, and there were moderate associations between *S* and *Z* (r(9998)=-0.77, p < 0.001) such that the minority group was more likely to benefit from treatment. For this group, treatment reduced risk of having an event and increased survival time. Higher values of the biomarker *X* were associated with a higher risk of an event (HR=1.98, 95% CI: [1.85, 2.12]). The simulated data were heavily right-censored to mirror the conditions of a prospective longitudinal study with finite follow-up period.

#### Treatment assignment under current risk-based and ITR approaches

The method used to calculate the pooled cohort equations (PCEs) from a given dataset was reproduced to reassign treatment under a risk assessment approach. A Cox PH model was fit for each *S* group to find a baseline 10-year survival rate. An individual “participant’s” risk score was calculated by estimating their probability of 10-year survival based on their value of *X* and their group’s baseline 10-year survival rate. If the 10-year risk of having an event was greater than 10%, the individual was assigned treatment *A*=1.

An ITR was estimated using *DWSurv*, an implementation of dynamic weighted survival modeling, a doubly robust optimal rule estimation procedure that can handle potentially right-censored survival time outcomes.^13^ The user specifies 4 models: (1) a “treatment-free” model, which is part of the accelerated failure time (AFT) model and represents the natural survival time in the absence of treatment, (2) a blip model, which is also part of the AFT model and represents the effect of the interaction between subject characteristics and treatment on the survival time, (3) the treatment model, which is a logistic regression used to estimate the effect of subject characteristics on the probability of treatment assignment, and (4), a censoring model, which is a logistic regression used to estimate the effect of subject characteristics on the probability of being censored. (1) and (2) are specified to estimate potential counterfactual survival times while (3) and (4) are used for balancing (i.e., inverse probability of treatment weighting), which aims to make treated and untreated groups comparable as they would be in a randomized controlled trial. We specified all four models using the interaction between the two available pieces of information: biomarker *X* and group *S. DWSurv* returned an ITR, which recommended a treatment decision for each subject based on the comparison of expected outcomes under treatment and no treatment on the absolute scale.

#### Outcomes

Since the true optimal treatment rule was known in the simulation study (recommend treatment to individuals with *Z*=0), we directly calculated the accuracy of each treatment assignment rule. We defined accuracy as the proportion of individuals in the sample treated optimally under the given rule. We also calculated this metric for each subgroup of *S* to determine whether each approach performed equally well across groups. The area under the curve (AUC) of the estimated Cox PH model was also calculated to confirm that this model worked well for predicting the risk of having an event.

### MESA Study

#### Study population

The Multiethnic Study of Atherosclerosis (MESA) is a multicenter, longitudinal cohort study of CVD risk factors, which recruited a socioeconomically diverse sample of adults, aged 45-84 years old, and free of clinical CVD in 2000-2002.^14^ The institutional review boards at all participating centers approved the study, and all participants gave informed consent. Participants were included in this analysis if they reported taking no antihypertensive medications at baseline. Enrollment and baseline examinations took place from 2000-2002; any participant with more than a 730-day duration between their baseline and second exam was excluded to minimize uncertainty about when treatment was initiated. Participants who experienced a CVD event or who were lost to follow-up before the second exam were excluded since survival time was measured from treatment initiation (Exam 2). In our main analysis, 3,452 participants met these inclusion criteria. Out of this group, 171 were missing data on key covariates listed below and were excluded from the analysis. The final sample included 3,281 participants.

#### Exposures

Participants who initiated any antihypertensives by their second exam, between 2002-2004, were considered treated, while those who did not initiate antihypertensives during this time were considered untreated.^15^ Treatment assignment was not randomized, and treatment adherence was ignored. This definition of treatment history is necessary for estimating a participant’s outcome under treatment vs no treatment, the basis of the ITR.

#### Covariates

Racial and ethnic group (Black, Chinese American, Hispanic, or White), gender (male or female), age, smoking status, and diabetes status were ascertained via questionnaire at the baseline examination. Total cholesterol and high-density lipoprotein (HDL) cholesterol were measured via blood samples following a 12-hour fast. Systolic blood pressure (SBP) was measured 3 times with participants seated. The mean of the second two measurements was recorded.^16^

#### Clinical outcomes

All participants were followed with regular phone contact every 9-12 months. The outcome of interest was any hard CVD event in a 10-year follow-up period, which could span from treatment assignment through 2012. Event-free survival time for each subject was counted from the first day antihypertensive use was reported. Information on these events were gathered from death certificates, hospital records, autopsy reports, patient interviews, or interviews administered to physicians, relatives, or friends, postmortem.^14^ Definition of a hard CVD event included myocardial infarction, resuscitated cardiac arrest, stroke (not transient ischemic attack), coronary heart disease death, and stroke death. Deaths that were unrelated to CVD were not counted as events. This outcome definition matched the one used in the PCE-based ASCVD risk score.^17^

#### Treatment assignment rule analysis

Treatment assignments based on risk scores versus an estimated ITR were compared. A 10-year ASCVD risk score was calculated for each subject at baseline by plugging their characteristics into the PCEs, using the published group-specific coefficients.^17^ According to 2017 ACC/AHA guidelines, those with a SBP of 130-139 mm Hg or a diastolic blood pressure (DBP) of 80-89 mm Hg (i.e., Stage 1 hypertension) with an estimated 10-year CVD risk of at least 10% should start antihypertensive therapy, and so should those with a SBP of at least 140 mm Hg or DBP 90 mm Hg (i.e., Stage 2 hypertension). This rule was used to decide treatment assignment under the risk score approach. An ITR was estimated using *DWSurv*.^13^ All and only variables that were used in the calculation of the 10-year ASCVD risk score were included in ITR estimation; it was most instructive to compare the two approaches to treatment recommendation using the same information. The ITR directly provided estimated optimal treatment assignments for each subject given the covariates listed above. The estimated ITR is found in Equation 1.

Treat if 1.398 + 0.001 ^*^ Age - 0.010 ^*^ Total Cholesterol - 0.009 ^*^ HDL + 0.009 ^*^ SBP + 0.824

^*^ *I*(Smoke) - 0.712 ^*^ *I*(Diabetes) + 2.072 ^*^ *I*(Black) + 1.509 ^*^ *I*(Hispanic)

+ 0.222 ^*^ *I*(Chinese American) - 1.191 ^*^ *I*(Male) + 0.047 ^*^ Age

^*^(Black or Hispanic) - 0.047 ^*^ Age ^*^ *I*(Male) + 0.006 ^*^ Total Cholesterol

^*^*I*(Black or Hispanic) + 0.007 ^*^ Total Cholesterol ^*^ *I*(Male) - 0.050 ^*^ HDL

^*^*I*(Black or Hispanic) + 0.037 ^*^ HDL ^*^ *I*(Male) - 0.028 ^*^ SBP

^*^*I*(Black or Hispanic) + 0.012 ^*^ SBP ^*^ *I*(Male) - 0.087 ^*^ *I*(Smoke) ^*^ *I*(Black)

– 0.471 ^*^ *I*(Smoke) ^*^ *I*(Male) - 0.077 ^*^ *I*(Diabetes) ^*^ *I*(Black or Hispanic)

+ 0.010 ^*^ *I*(Diabetes) ^*^ *I*(Male) > 0

Equation 1: Individualized treatment rule estimated from the MESA sample. A patient’s characteristics are plugged in, and treatment recommendations are made based on whether the value falls above or below 0. “I()” represents an indicator function that is equal to 1 when the item in parentheses is true and 0 otherwise.

#### Treatment assignment rule outcomes

A Chi-square test of independence was performed to evaluate whether treatment decisions and racial and ethnic category were significantly associated. Chi-square tests and t-tests were performed to identify statistically significant differences in demographic variables between the group that is only treated under the ITR (n = 1,570) and the group that is only treated under the risk assessment approach (n = 254).

#### Sensitivity Analyses

Four sensitivity analyses were conducted. The first included subjects who took longer than two years to attend their second MESA exam. The second compared the ITR estimated in the main analysis to a risk score (Cox PH model) estimated from the MESA sample, which could be different from the score based on the PCEs. The third compared the risk score-based rule to the ITR that was estimated when the treatment initiation window shifted to consider the fourth MESA exam as the baseline exam and considered subjects who reported antihypertensive use at Exam 5 as treated. The fourth compared the risk assessment rule to the ITR that was estimated from the sub-sample that reported high blood pressure at baseline. The fifth compared the ITR estimated in the main analysis to a rule that used the PREVENT equations to calculate risk scores.^6^ Details of these analyses are found in the Supplement.

## Results

### Simulation Study

Under the risk prediction model, 44% of “subjects” were treated optimally (i.e., in agreement with their unobserved value of *Z*), while under the ITR, 90% were treated optimally. These accuracy metrics were stratified, as shown in Figure 2, to show rates of correct vs incorrect treatment decisions across *S* groups. Using the risk score algorithm, 90% of subjects in the “minority” group were incorrectly untreated, while 42% of subjects in the “majority” group were incorrectly treated (overtreatment). Using the ITR, incorrect treatment decisions were substantially reduced in both groups, with 10% of the “minority” group incorrectly treated and 10% of the “majority” group incorrectly untreated. The risk assessment method worked well for predicting events (shown in the Supplement) but did not assign treatments optimally.

**Figure 2:**
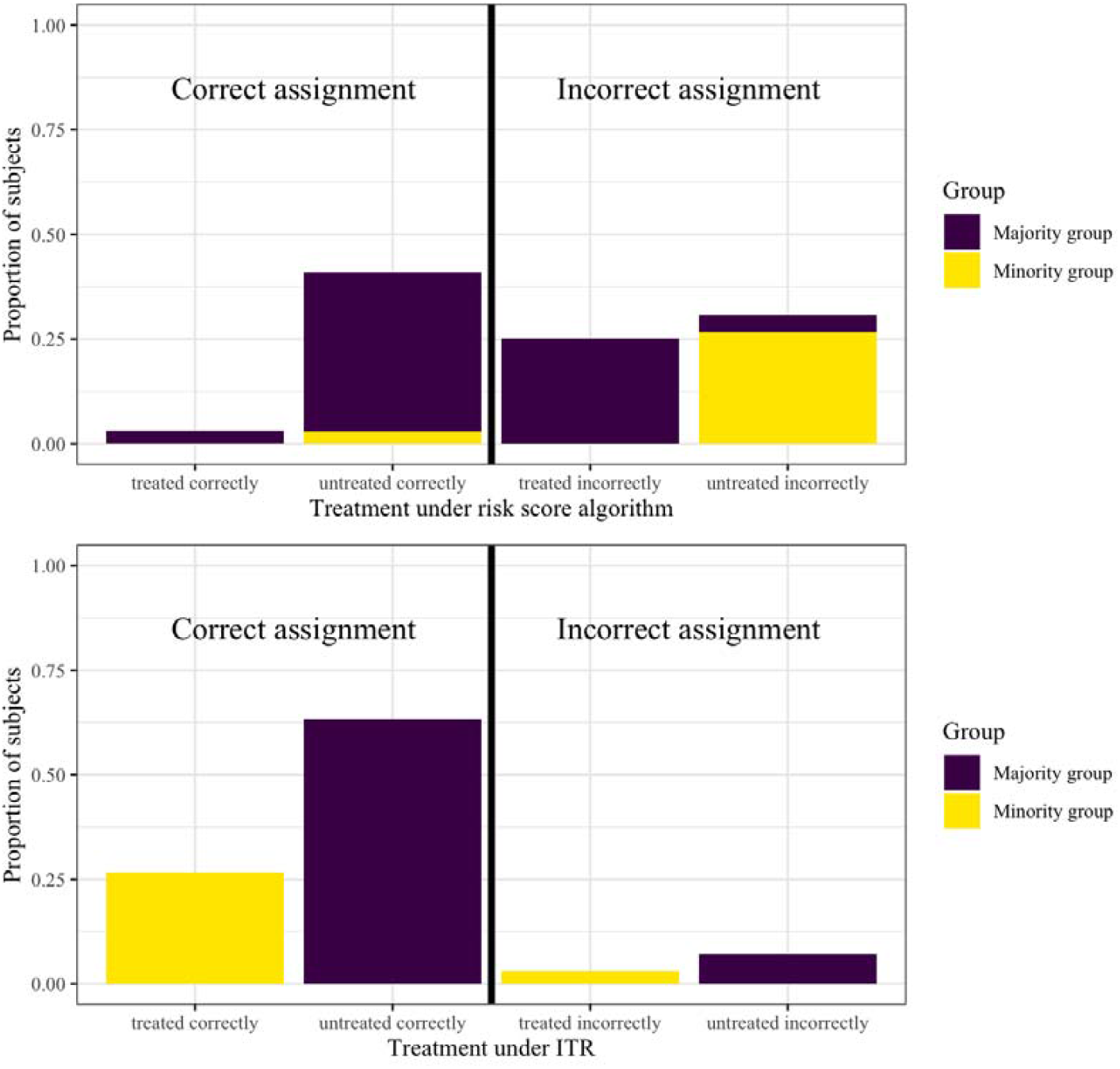
Treatment accuracy by group for each method in simulation study

### MESA

Demographics and key characteristics of the sample are summarized in Table 1.

**Table 1:** Demographic information of MESA sample meeting enrollment criteria at baseline.

| Baseline characteristic | Reported antihypertensive use<br>(n=376) | Did not report antihypertensive<br>use (n=2,905) |
| --- | --- | --- |
| Age, mean (SD) | 64.6 (9.9) | 60.0 (10.0) |
| Gender, n (%) |  |  |
| Male | 187 (50%) | 1,422 (49%) |
| Female | 189 (50%) | 1,483 (51%) |
| Racial and ethnic group, n (%) |  |  |
| Black | 122 (32%) | 623 (21%) |
| Chinese American | 39 (10%) | 406 (14%) |
| Hispanic | 79 (21%) | 649 (22%) |
| White | 136 (36%) | 1,227 (42%) |
| Total cholesterol, mean (SD) | 194.6 (35.3) | 196.6 (34.8) |
| HDL cholesterol, mean (SD) | 49.6 (14.0) | 51.6 (15.1) |
| Systolic blood pressure, mean (SD) | 139.2 (21.8) | 119.6 (18.0) |
| Smoking status, n (%) |  |  |
| Smokes | 46 (12%) | 412 (14%) |
| Does not smoke | 330 (88%) | 2,493 (86%) |
| Diabetes status, n (%) |  |  |
| Has diabetes | 62 (16%) | 159 (5.5%) |
| Does not have diabetes | 314 (84%) | 2,746 (94.5%) |
| Hard CVD event, n (%) |  |  |
| Experienced event | 44 (12%) | 138 (4.8%) |
| Did not experience event | 332 (88%) | 2,767 (95.2%) |

The risk assessment approach recommended treatment to 861 subjects while the ITR recommended treatment to 2,177 subjects. Only 607 subjects were recommended treatment under both rules. Treatment assignment rates for all methods are shown in Table 2, where the ITR recommended higher treatment rates for all racial and ethnic groups when compared to the recommendations from the risk score approach.

**Table 2:** Suggested treatment rates at baseline (Exam 1) for each race under different treatment assignment scenarios.

| Racial and ethnic group | # to be treated under ACC/AHA guideline | # to be treated under ITR | # treated (observed) |
| --- | --- | --- | --- |
| Black (745) | 238 (32%) | 513 (69%) | 122 (16%) |
| Chinese American (445) | 102 (23%) | 359 (81%) | 39 (8.8%) |
| Hispanic (728) | 195 (27%) | 445 (61%) | 79 (11%) |
| White (1,363) | 326 (24%) | 860 (63%) | 136 (10%) |

Under the ITR, Black and Chinese American participants were more likely to be recommended treatment than White participants, while Hispanic participants were less likely to be recommended treatment when compared to White participants. Under the ACC/AHA guidelines, Black and Hispanic participants were more likely to be recommended treatment than White and Chinese American subjects, and the overall treatment rate was much lower for all groups. The group differences in treatment rates also varied somewhat when comparing treatment under the two rules, though the relative treatment rate differences were less pronounced than the absolute differences for all groups. A Chi-square test for independence of racial and ethnic group and treatment rule rejected the null hypothesis that there is no association between the variables at the alpha = 0.01 level (X^2^ = 28.75, d*f* = 6, *p* < 0.001). Furthermore, we calculated the magnitude of the contribution of each cell towards the Chi-square statistic by looking at its residual. From this, we saw that the largest contributions to the Chi-square statistic came from the difference in treatment rates for Black subjects that were truly observed on treatment vs recommended treatment under the ITR and from the large difference in treatment assignment rates suggested under each rule for Chinese Americans. Residuals are shown in Table 3. Table 4 describes the differences in the distribution of subjects that were suggested treatment under only one rule and not the other. Subjects were different in all the listed covariates, e.g., those treated under the ITR but not the ACC/AHA guidelines were likely to be younger, male, and healthier according to various measures (lower SBP and cholesterol).

**Table 3:** Residual contributions to Chi-square statistic in test for independence.

| Racial and ethnic group | Residual contribution under ACC/AHA guideline | Residual contribution under ITR | Residual contribution under observed data |
| --- | --- | --- | --- |
| Black (745) | 5.0 | **11.9** | **24.2** |
| Chinese American (445) | **16.0** | **17.6** | **16.3** |
| Hispanic (728) | 3.6 | 1.4 | < 0.0 |
| White (1,363) | 0.6 | 1.2 | 2.2 |

**Table 4:**
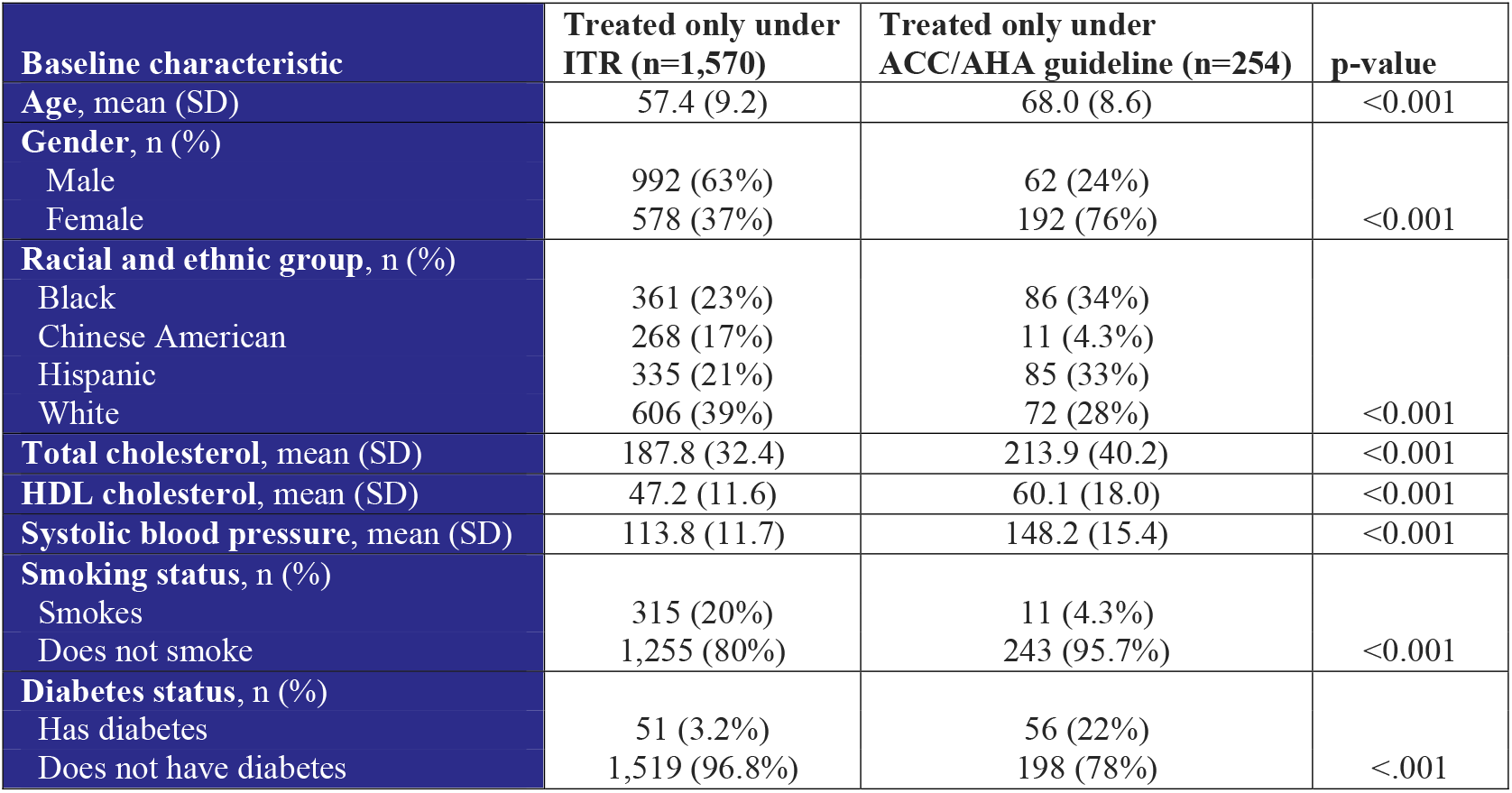
Frequencies and averages for demographic characteristics of subgroups treated only under the ITR or only under the risk assessment algorithm. Given p-values refer to either a Chi-square or t-test given whether the variable is categorical or continuous, respectively. All p-values are Bonferroni adjusted to account for multiple comparisons.

The sensitivity analyses results showed that relaxing the enrollment criterion to include subjects with more than two years between their baseline and exam 2 visits (Analysis 1) did not yield substantively different results, neither did updating risk scores by re-estimating PCEs from the MESA sample (Analysis 2). When more recent exam data were used for analysis (Analysis 3), the observed treatment rates increased, but the ITR and ACC/AHA recommendations did not change drastically. Finally, restricting the sample to subjects with hypertension at baseline (Analysis 4) did produce substantially different treatment recommendation results.

## Discussion

This study contrasted the standard approach to treatment recommendations for hypertension, based on thresholding a global risk score, with an approach that used an estimated ITR. The relevant concepts were illustrated with a simulation to emphasize the technical differences between the two decision-making paradigms in the face of treatment effect heterogeneity. The study then investigated the use of antihypertensive medications in MESA; an ITR was compared with a treatment rule based on ACC/AHA guidelines using the PCE-derived risk score.^17^ When models are correctly specified and requisite design assumptions are met to make conditional treatment effects identifiable, we expect the ITR to recommend treatment optimally, i.e., in a way that maximizes mean event-free survival time.

The simulation study provided “in-principle” evidence that ITRs may make substantially different, and better, treatment recommendations compared to an approach based on risk prediction in the presence of treatment heterogeneity, and that treatment rates may vary across demographic groups when group membership is associated with relevant variables. The key benefit of using synthetic data here was that the truly “optimal” treatment decision was known so error rates could be calculated. In the data generated, simply moving from a risk prediction approach to estimating ITRs drastically reduced disparities in treatment and error rates across groups. The analysis of MESA data suggested that the recommendation of antihypertensives to prevent cardiovascular events may differ substantially when comparing an ITR model with a risk assessment algorithm. The difference in treatment assignment rates under these different approaches varied across racial and ethnic groups, with the ITR recommending treatment to more patients overall.

### Limitations

The primary limitation of the MESA analysis is the inability to validate which treatment assignment rule is optimal. While the ITR attempts to uncover the optimal rule, this relies on several assumptions and correct specification of at least some of the models used in the estimation procedure. While the simulation underscores the key conceptual point – that an ITR may be preferable from both “optimality” and “equity” perspectives – our analysis of the real MESA data can only support the more measured conclusion that the two approaches to treatment recommendation may produce substantially different results. Whereas future research may propose an estimated ITR based on high-quality (observational or trial) data, the present study provides the first empirical support to the hypothesis that informing treatment decisions by ITR versus risk prediction may make a difference to which patients are recommended treatment.

Although the MESA sample used in this study is more racially and ethnically diverse than many other cohort studies, relatively low numbers of Chinese Americans in various substrata lead to substantially different treatment recommendation rates for this group between the main and sensitivity analyses.

Results from this analysis could be biased by variation in treatment start dates. A participant’s treatment initiation time was identified with their Exam 2 date. Even if a participant began antihypertensive treatment immediately after their baseline exam, the data would not reflect this – only whether they were reportedly taking antihypertensives by Exam 2. Therefore, an association between treatment assignment and true treatment initiation time (rather than the operationalized time from the data), could positively bias the estimated treatment effect on event-free survival if participants taking antihypertensives disproportionately had later Exam 2 dates or bias the effect towards the null if these participants had earlier Exam 2 dates. This bias would negatively impact the accuracy of the estimated ITR.

### Future Directions

In this study, we used the same set of covariates in the ITR as were used in the PCE-based risk score to facilitate fair comparison. It is possible that additional variables that are available in MESA data but not included in our analysis could improve the performance of the ITR. Future analyses could explore the benefit gained from including other clinical variables and social determinants of health.

ITRs may be estimated to recommend one form of treatment over another. It may be of interest for clinicians in this setting to know whether one class of antihypertensive medications, when compared to another class, is expected to prolong the survival time of a patient with specific characteristics.^18^ Future studies may consider such questions, which may be even more pertinent to clinical practice than the one explored here.

## Data Availability

MESA ancillary study PIs are restricted from directly sharing any MESA contract-derived data, including the MESA ID, age, sex, race/ethnicity, field center, or other data, without MESA permission. Individual participant data after de-identification can be made available to investigators who have an approved study proposal by the MESA coordinating center. This includes study protocol and data dictionaries. These are only available following this approval. Simulation code and data can be found at https://github.com/dmalinsk/TreatmentRules.

## Acknowledgements

This research was partially supported by the Calderone Health Equity Award from the Columbia Mailman School of Public Health. The authors thank the other investigators, the staff, and the participants of the MESA study for their valuable contributions. A full list of participating MESA investigators and institutions can be found at http://www.mesa-nhlbi.org.

## Sources of Funding

This research was supported by contracts 75N92020D00001, HHSN268201500003I, N01-HC-95159, 75N92020D00005, N01-HC-95160, 75N92020D00002, N01-HC-95161, 75N92020D00003, N01-HC-95162, 75N92020D00006, N01-HC-95163, 75N92020D00004, N01-HC-95164, 75N92020D00007, N01-HC-95165, N01-HC-95166, N01-HC-95167, N01-HC-95168 and N01-HC-95169 from the National Heart, Lung, and Blood Institute, and by grants UL1-TR-000040, UL1-TR-001079, and UL1-TR-001420 from the National Center for Advancing Translational Sciences (NCATS).

## Notes

### Competing Interest Statement

The authors have declared no competing interest.

### Author Declarations

The institutional review boards at all participating centers approved the study, and all participants gave informed consent.

## References

1. Khan SS, Coresh J, Pencina MJ, Ndumele CE, Rangaswami J, Chow SL, Palaniappan LP, Sperling LS, Virani SS, Ho JE, Neeland IJ, Tuttle KR, Rajgopal Singh R, Elkind MSV, Lloyd-Jones DM, on behalf of the American Heart Association. Novel Prediction Equations for Absolute Risk Assessment of Total Cardiovascular Disease Incorporating Cardiovascular-Kidney-Metabolic Health: A Scientific Statement From the American Heart Association. Circulation. 2023;148:1982–2004.

2. Recommendation: Statin Use for the Primary Prevention of Cardiovascular Disease in Adults: Preventive Medication | United States Preventive Services Taskforce [Internet]. [cited 2024 Jul 17];Available from: https://www.uspreventiveservicestaskforce.org/uspstf/recommendation/statin-use-in-adults-preventive-medication

3. Goff DC, Lloyd-Jones DM, Bennett G, Coady S, D’Agostino RB, Gibbons R, Greenland P, Lackland DT, Levy D, O’Donnell CJ, Robinson JG, Schwartz JS, Shero ST, Smith SC, Sorlie P, Stone NJ, Wilson PWF, Jordan HS, Nevo L, Wnek J, Anderson JL, Halperin JL, Albert NM, Bozkurt B, Brindis RG, Curtis LH, DeMets D, Hochman JS, Kovacs RJ, Ohman EM, Pressler SJ, Sellke FW, Shen W-K, Smith SC, Tomaselli GF, American College of Cardiology/American Heart Association Task Force on Practice Guidelines. 2013 ACC/AHA guideline on the assessment of cardiovascular risk: a report of the American College of Cardiology/American Heart Association Task Force on Practice Guidelines. Circulation. 2014;129:S49–73.

4. Whelton PK, Carey RM, Aronow WS, Casey DE, Collins KJ, Dennison Himmelfarb C, DePalma SM, Gidding S, Jamerson KA, Jones DW, MacLaughlin EJ, Muntner P, Ovbiagele B, Smith SC, Spencer CC, Stafford RS, Taler SJ, Thomas RJ, Williams KA, Williamson JD, Wright JT. 2017 ACC/AHA/AAPA/ABC/ACPM/AGS/APhA/ASH/ASPC/NMA/PCNA Guideline for the Prevention, Detection, Evaluation, and Management of High Blood Pressure in Adults: Executive Summary: A Report of the American College of Cardiology/American Heart Association Task Force on Clinical Practice Guidelines. Hypertension. 2018;71:1269–1324.

5. Lloyd-Jones DM, Braun LT, Ndumele CE, Smith SC, Sperling LS, Virani SS, Blumenthal RS. Use of Risk Assessment Tools to Guide Decision-Making in the Primary Prevention of Atherosclerotic Cardiovascular Disease: A Special Report From the American Heart Association and American College of Cardiology. J Am Coll Cardiol. 2019;73:3153–3167.

6. Khan SS, Matsushita K, Sang Y, Ballew SH, Grams ME, Surapaneni A, Blaha MJ, Carson AP, Chang AR, Ciemins E, Go AS, Gutierrez OM, Hwang S-J, Jassal SK, Kovesdy CP, Lloyd-Jones DM, Shlipak MG, Palaniappan LP, Sperling L, Virani SS, Tuttle K, Neeland IJ, Chow SL, Rangaswami J, Pencina MJ, Ndumele CE, Coresh J, for the Chronic Kidney Disease Prognosis Consortium and the American Heart Association Cardiovascular-Kidney-Metabolic Science Advisory Group. Development and Validation of the American Heart Association’s PREVENT Equations. Circulation. 2024;149:430–449.

7. Paulus J, Kent D. Predictably unequal: understanding and addressing concerns that algorithmic clinical prediction may increase health disparities. npj Digital Medicine. 2020;3:99.

8. Qian M, Murphy SA. Performance guarantees for individualized treatment rules. The Annals of Statistics. 2011;39:1180–1210.

9. Murphy SA. Optimal dynamic treatment regimes. Journal of the Royal Statistical Society Series B. 2003;65:331–355.

10. Fahmi AM, Elewa H, El Jilany I. Warfarin dosing strategies evolution and its progress in the era of precision medicine, a narrative review. Int J Clin Pharm. 2022;44:599–607.

11. Kent DM, Paulus JK, van Klaveren D, D’Agostino R, Goodman S, Hayward R, Ioannidis JPA, Patrick-Lake B, Morton S, Pencina M, Raman G, Ross JS, Selker HP, Varadhan R, Vickers A, Wong JB, Steyerberg EW. The Predictive Approaches to Treatment effect Heterogeneity (PATH) Statement. Ann Intern Med. 2020;172:35–45.

12. Inoue K, Athey S, Tsugawa Y. Machine-learning-based high-benefit approach versus conventional high-risk approach in blood pressure management. Int J Epidemiol. 2023;52:1243–1256.

13. Simoneau G, Moodie E, Wallace M, Platt R. Optimal dynamic treatment regimes with survival endpoints: introducing DWSurv in the R package DTRreg. Journal of Statistical Computation and Simulation. 2020;90:1–18.

14. Bild DE, Bluemke DA, Burke GL, Detrano R, Diez Roux AV, Folsom AR, Greenland P, Jacob DR, Kronmal R, Liu K, Nelson JC, O’Leary D, Saad MF, Shea S, Szklo M, Tracy RP. Multi-Ethnic Study of Atherosclerosis: objectives and design. Am J Epidemiol. 2002;156:871–881.

15. Hernán MA, Robins JM. Using Big Data to Emulate a Target Trial When a Randomized Trial Is Not Available. Am J Epidemiol. 2016;183:758–764.

16. Bild DE, Detrano R, Peterson D, Guerci A, Liu K, Shahar E, Ouyang P, Jackson S, Saad MF. Ethnic differences in coronary calcification: the Multi-Ethnic Study of Atherosclerosis (MESA). Circulation. 2005;111:1313–1320.

17. Muntner P, Colantonio LD, Cushman M, Goff DC, Howard G, Howard VJ, Kissela B, Levitan EB, Lloyd-Jones DM, Safford MM. Validation of the atherosclerotic cardiovascular disease Pooled Cohort risk equations. JAMA. 2014;311:1406–1415.

18. Cushman WC, Reda DJ, Perry HM, Williams D, Abdellatif M, Materson BJ. Regional and racial differences in response to antihypertensive medication use in a randomized controlled trial of men with hypertension in the United States. Department of Veterans Affairs Cooperative Study Group on Antihypertensive Agents. Arch Intern Med. 2000;160:825–831.

