## Supplement for "A health equity perspective on data-driven treatment decisions in cardiovascular care: risk assessments versus individualized treatment rules"

Figure 3 shows the receiver operating characteristic (ROC) curve for the risk assessment method. This curve shows the sensitivity/specificity of the model for predicting events and has an AUC of 0.73.

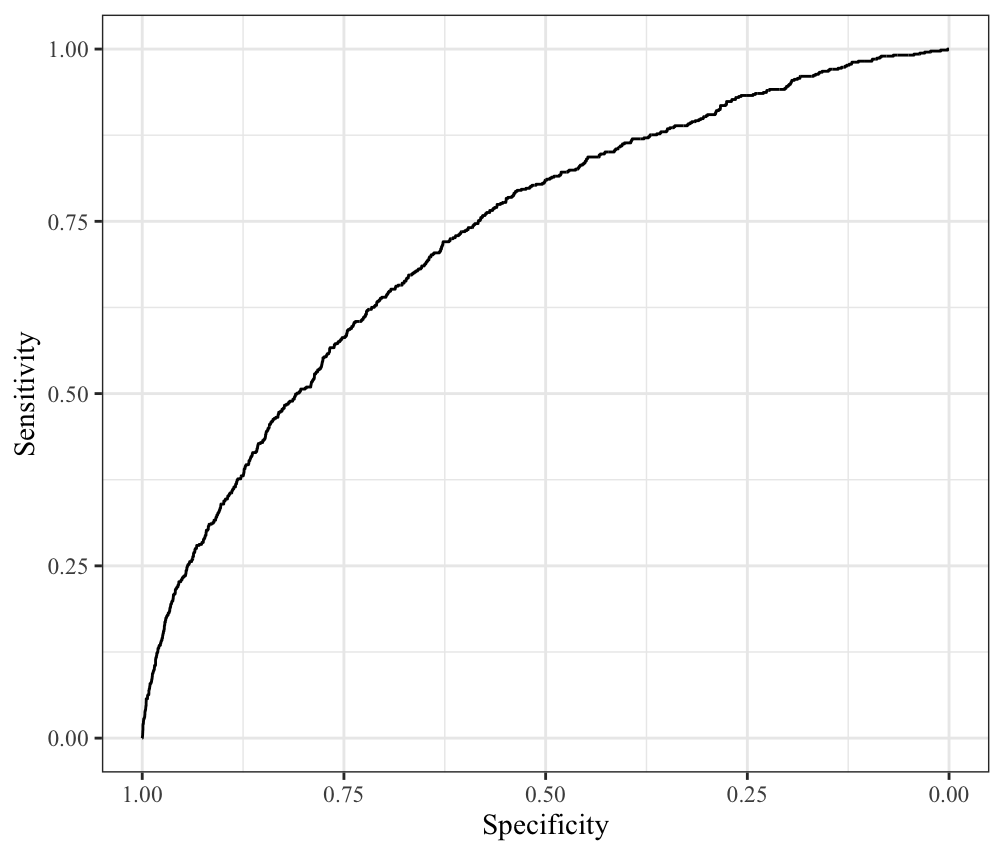

Figure 3: ROC curve for risk score method

**Sensitivity Analyses**

**Analysis 1: No restriction on treatment assignment window.** The first sensitivity analysis includes subjects who take longer than two years to attend their second MESA exam, while the main analysis only includes those with a duration between the two exams fewer than 730 days. This allows for a slightly larger sample, which makes results generalizable to a wider population. However, there is an increased chance for biased results, given a subject’s actual time under treatment may be over 2 years apart from the time treatment is coded as beginning for the purposes of the analysis. In this analysis, the risk assessment rule and ITR are estimated and applied in the same manner as in the main analysis, but on a larger sample (n = 3,710). Results are found in Table 5. Results of this sensitivity analysis are similar to the results of our main analysis.

Table 5: Treatment recommendations when the sample is not limited by time between baseline and Exam 2.

| **Racial/ethnic group** | **# to be treated under ACC/AHA guideline** | **# to be treated under ITR** | **# treated (observed)** |
| --- | --- | --- | --- |
| White (1,535) | 358 (23%) | 832 (54%) | 157 (10%) |
| Black (816) | 254 (31%) | 521 (64%) | 135 (17%) |
| Hispanic (853) | 215 (25%) | 543 (64%) | 95 (11%) |
| Chinese American (506) | 112 (22%) | 397 (78%) | 49 (9.7%) |

**Analysis 2: Updated risk score.** The second sensitivity analysis re-estimates the PCEs used to assign a risk score to each subject in the sample by fitting Cox PH models on the present sample, rather than assigning risk scores based on coefficients estimated from previous studies. In this analysis, only the proportions of subjects treated under the ACC/AHA guideline change, while the estimated ITR and the given sample remain the same. Results are found in Table 6. Using this updated risk score changes the result only marginally.

Table 6: Treatment recommendations when risk scores are estimated based on Cox regressions fit on the current MESA sample.

| **Racial/ethnic group** | **# to be treated under ACC/AHA guideline** | **# to be treated under ITR** | **# treated (observed)** |
| --- | --- | --- | --- |
| White (1,363) | 334 (25%) | 860 (63%) | 136 (10%) |
| Black (745) | 228 (31%) | 513 (69%) | 122 (16%) |
| Hispanic (728) | 197 (27%) | 445 (61%) | 79 (11%) |
| Chinese American (445) | 99 (22%) | 359 (81%) | 39 (8.8%) |

**Analysis 3: Considering Exam 4 as baseline.** In this analysis, more current data is used to better reflect present practices in treatment assignment. Specifically, Exam 4, which took place between 2005 and 2007, is treated as the baseline exam. Those who report antihypertensive treatment by Exam 5, which took place between 2010 and 2012, are considered as assigned to treatment. The main analysis techniques are applied with respect to this new timeline and sample (n = 2,262) to find treatment assignment rates under the ACC/AHA guideline and under a newly estimated ITR. Results are found in Table 7. The results show that the number to be treated under ACC/AHA guidelines and the number who are treated in reality are similar. This may reflect the adaptation of clinicians to the guidelines. The ITR continues to recommend higher treatment rates than the risk-based guideline.

Table 7: Treatment recommendations when treatment assignment is defined as antihypertensive initiation between baseline and Exam 5.

| **Racial/ethnic group** | **# to be treated under ACC/AHA guideline** | **# to be treated under ITR** | **# treated (observed)** |
| --- | --- | --- | --- |
| White (1,005) | 177 (18%) | 608 (60%) | 198 (20%) |
| Black (444) | 127 (29%) | 294 (66%) | 137 (31%) |
| Hispanic (511) | 111 (22%) | 278 (54%) | 128 (25%) |
| Chinese American (302) | 58 (19%) | 181 (60%) | 68 (23%) |

**Analysis 4: Restricting sample to those with high blood pressure.** The fourth sensitivity analysis compares the risk assessment rule to the ITR that are estimated from the sample that experiences high blood pressure (i.e., systolic blood pressure greater than 130 mm Hg or diastolic blood pressure greater than 80 mm Hg) at baseline (n = 1,188). Under the ACC/AHA guideline, a subject from this sample should be assigned treatment if their ASCVD risk score is greater than 10% or if they have stage 2 hypertension (i.e., systolic blood pressure greater than 140 mm Hg or diastolic blood pressure greater than 90 mm Hg). Results are found in Table 8. For this sample, the ITR recommends treatment to fewer Black and Hispanic subjects, as it is more heavily influenced by biomarkers and demographic factors other than blood pressure level.

Table 8: Treatment recommendations when sample only includes those with high blood pressure at baseline.

| **Racial/ethnic group** | **# to be treated under ACC/AHA guideline** | **# to be treated under ITR** | **# treated (observed)** |
| --- | --- | --- | --- |
| White (447) | 326 (73%) | 388 (87%) | 91 (20%) |
| Black (332) | 238 (72%) | 183 (55%) | 90 (27%) |
| Hispanic (258) | 195 (76%) | 114 (44%) | 51 (20%) |
| Chinese American (151) | 102 (68%) | 149 (99%) | 29 (19%) |

**Analysis 5: PREVENT-based risk score.** The fifth sensitivity analysis calculates baseline 10-year ASCVD risk using the PREVENT equations, rather than the PCEs. In this analysis, only the proportions of subjects treated under the ACC/AHA guideline change, while the estimated ITR and the given sample remain the same. Results are found in Table 9. Under the rule using the PREVENT-based risk score, each racial/ethnic group is recommended treatment at a lower rate when compared to the PCE-based rule and the ITR. These results agree with research suggesting that using PREVENT-based scores along with ACC/AHA guidelines might reduce the number of patients eligible for statins and antihypertensive medication.^1^ Although a PREVENT calculator is publicly available, the most recent ACA/AHA guidelines still use the PCE-based scores for clinical decision-making.

Table 9: Treatment recommendations when risk scores are estimated based on the PREVENT equations.

| **Racial/ethnic group** | **# to be treated under ACC/AHA guideline** | **# to be treated under ITR** | **# treated (observed)** |
| --- | --- | --- | --- |
| White (1,363) | 251 (18%) | 860 (63%) | 136 (10%) |
| Black (745) | 198 (27%) | 513 (69%) | 122 (16%) |
| Hispanic (728) | 149 (20%) | 445 (61%) | 79 (11%) |
| Chinese American (445) | 85 (19%) | 359 (81%) | 39 (8.8%) |

**References**

1.	Anderson TS, Wilson LM, Sussman JB. Atherosclerotic Cardiovascular Disease Risk Estimates Using the Predicting Risk of Cardiovascular Disease Events Equations. *JAMA Internal Medicine*. 2024;184(8):963-970. doi:10.1001/jamainternmed.2024.1302
